# Separating the effects of immune waning and viral evolution on COVID-19 vaccine effectiveness reveals improved effectiveness of updated vaccines

**DOI:** 10.64898/2026.09.14.26363077

**Authors:** Wang Jin, Ainslie Mitchell, Eva Stadler, Charles S.P. Foster, Timothy E. Schlub, Miles P. Davenport, Deborah Cromer

**Affiliations:** Kirby Institute, University of New South Wales, Sydney, New South Wales, Australia; School of Population Health, Faculty of Medicine and Health, University of New South Wales, Sydney, New South Wales, Australia; School of Biomedical Sciences, Faculty of Medicine and Health, University of New South Wales, Sydney, New South Wales, Australia; Sydney School of Public Health, Faculty of Medicine and Health, The University of Sydney, Sydney, New South Wales, Australia

## Abstract

The continued evolution of SARS-CoV-2 has led to regular updates of the COVID-19 vaccine immunogen. However, the exact benefits of these updates for vaccine effectiveness in a real world context are difficult to evaluate. We performed a comprehensive meta-analysis of booster vaccine effectiveness in studies identified through the ViewHub systematic review. We integrated evidence from 44 studies reporting on effectiveness of mRNA booster vaccines containing XBB.1.5, JN.1 and KP.2 immunogens to assess the combined impact of booster immunogen, time since vaccination and calendar time at which a study was conducted. We estimate that the relative risk of severe disease increases by 9% [95%CI 7.4-10.5] each month following booster administration, likely due to antibody waning, and by an additional 2.4% [95%CI 1.2-3.6] for each calendar month, likely due to antigenic drift. Our findings show that COVID-19 booster vaccine effectiveness declines both with time after vaccination as well as calendar time since the vaccine immunogen emerged. This reinforces the need for regularly and timely vaccine immunogen updates.

## Introduction

Since its emergence in late 2019, severe acute respiratory syndrome coronavirus 2 (SARS-CoV-2) has transitioned from a pandemic pathogen to an endemic respiratory virus characterised by ongoing antigenic drift allowing it to escape existing immune responses and leading to periodic replacement of the circulating variant. This has led to multiple recommendations for updates to the immunogen contained within the Coronavirus Disease 2019 (COVID-19) vaccine^1-3^, to improve vaccine-induced immunity and protection against current variants^4,5^.

While there is clear evidence that updating the vaccine immunogen increases antibody responses against more recent variants^6^, there is more limited evidence that this concomitantly increases vaccine effectiveness.

Given the potential ongoing improvement to both cellular responses and the broadening of antibody responses, it is possible that the impact of antigenic drift on vaccine effectiveness is no longer as strong as it once was. Understanding the relationship between the immunogen contained within a vaccine, its effectiveness and how this changes due to antigenic drift is now of critical importance. Such information will inform decisions regarding vaccine strain selection, vaccine update frequency and optimal booster scheduling.

In this work, we set out to determine how COVID-19 vaccine effectiveness has changed over time and in response to vaccine updates. We utilised data from the ViewHub systematic review database^7^, which aggregated data on vaccine effectiveness following a range of booster immunogens. We asked whether vaccine effectiveness decreases as the circulating variant evolves, and if so, whether the magnitude of this decrease can be quantified. The results of this work will not only significantly impact decisions regarding COVID-19 vaccine updates but will also present a methodological advancement that can be applied to other evolving pathogens requiring ongoing vaccine updates, such as influenza.

## Results

### Data Identified using ViewHub Systematic Review

We identified data from 44 different studies (22 test-negative case-control studies and 22 cohort studies). Of these, 33 reported on vaccine effectiveness after boosting with an XBB.1.5 containing vaccine, 4 after a JN.1 vaccine, 5 after a KP.2 vaccine and 4 after either a JN.1 or KP.2 vaccine (combined cohort) (Figure 1A).

**Figure 1.**
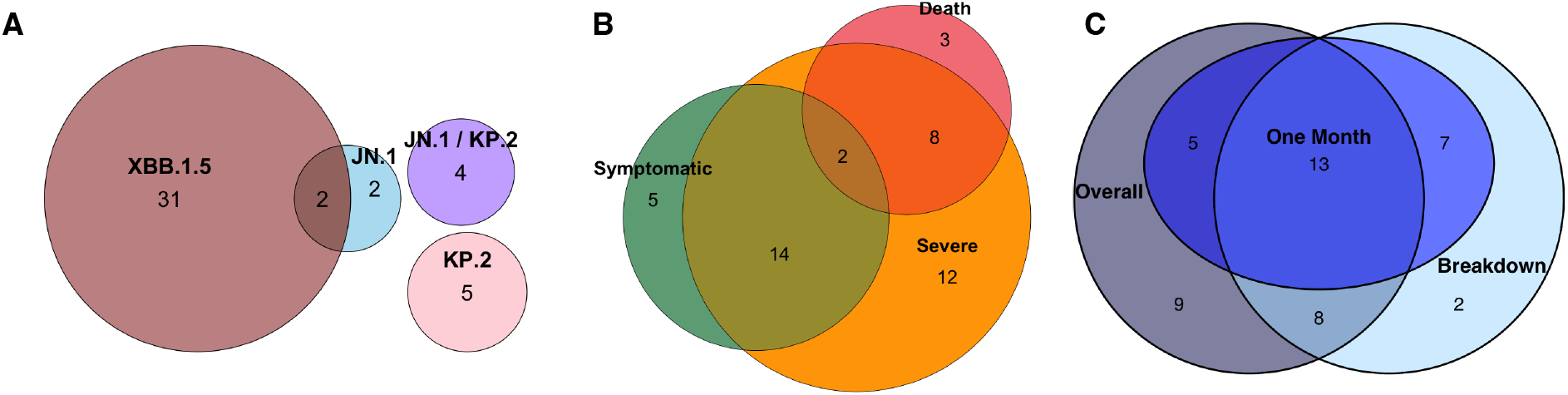
Venn Diagram of Studies showing the numbers of studies by (A) immunogen contained within the administered booster, (B) disease outcome reported and (C) time period over which vaccine effectiveness was reported.

The studies reported on vaccine effectiveness against symptomatic disease (21 studies), severe disease (36 studies) and death (13 studies) (Figure 1B). All except five of the studies reported on vaccine effectiveness in an adult only, immunocompetent population, with the remaining studies reporting on vaccine effectiveness in an immunocompetent population that included both children and adults.

Most of the studies (35/44) provided an overall vaccine effectiveness result for the duration of the study, while some studies (30/44) broke down the vaccine effectiveness by the time since vaccination. Nine studies did not provide an overall vaccine effectiveness estimate, and only provided VE estimates broken down by time intervals (Figure 1C). In addition, 25 studies provided a VE estimate at around one month following booster vaccination. (i.e. median time between 14 and 45 days following boosting, and maximum time < 60 days after boosting) (Figure 1C).

### Do vaccine updates improve vaccine effectiveness?

We first compared vaccine effectiveness following XBB.1.5 and JN.1/KP.2 vaccination. Across all studies we found no evidence that the newer JN.1/KP.2 immunogens had higher effectiveness than the XBB.1.5 vaccines when considering VE estimates either over the entire study period or within the first month following booster administration (Figure 2A, B; p=0.95 and p=0.14 for the booster immunogen variable in a meta-analytic mixed-effects model, respectively). At face value, this suggests that updating the vaccine immunogen provides little benefit.

**Figure 2.**
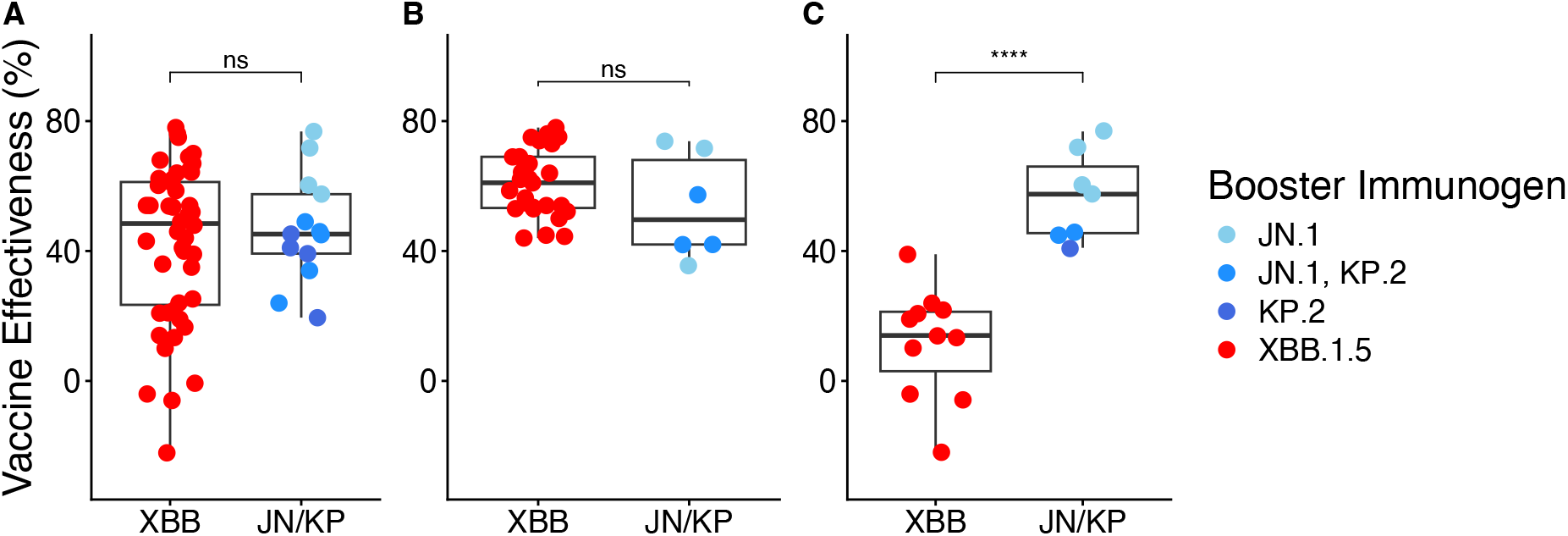
XBB vs JN/KP booster effectiveness against severe disease either (A) over the entire study period in all studies (B) over the first month since booster administration in all studies or (C) over the entire study period in studies conducted wholly between 1^st^ June 2024 and 31^st^ January 2025. Significance values shown are for unpaired t-tests, **** signifies p <= 0.0001.

However, studies evaluating booster vaccines containing XBB.1.5 and JN.1/KP.2 were conducted at different calendar times and in the context of different circulating variants. To minimise this confounding, we identified studies reporting vaccine effectiveness against severe disease that were conducted during a period (1^st^ June 2024 to 31^st^ January 2025) when both XBB.1.5 and JN.1/KP.2 vaccines were in use. Five studies met these criteria, of which 1 study reported on vaccine effectiveness (VE) following an XBB.1.5 booster (in 11 population groups) and 4 studies reported on vaccine effectiveness following a JN.1 and / or KP.2 booster (in 7 population groups).

Within this restricted dataset, effectiveness against severe disease was significantly higher following JN.1/KP.2 boosting than following XBB.1.5 boosting (median effectiveness against severe disease was 57.5% vs 14%; p=0.0019, for the booster immunogen variable in a meta-analytic mixed-effects model, Figure 2C).

We note that the fact that there was only 1 study reporting on effectiveness of an XBB.1.5 booster vaccine during this period highlights the difficulty of making such a direct comparison. Although we cannot rule out study specific effects, we note that the XBB.1.5 study reported vaccine effectiveness in cohorts from 6 different European countries across 2 different age groups, generating the 11 separate estimates of effectiveness against severe disease following XBB.1.5 vaccination shown in Figure 2C. The consistency across populations provides reassurance that the observed difference is not driven by a single narrowly defined cohort, however additional comparative studies would strengthen this finding.

### Is vaccine effectiveness impacted by the time at which a study is conducted?

Given the difference in reported vaccine effectiveness following XBB.1.5 boosting in either all studies (Figure 2A) or studies conducted at the later end of the period during which XBB.1.5 vaccines were being used (Figure 2C), we investigated whether the calendar time at which a study was conducted (as a proxy for antigenic drift) was associated with the vaccine effectiveness reported in the study.

To minimise the potential confounding effects of antibody waning and study duration, we considered only vaccine effectiveness estimates in the first month after boosting (see Methods). Where possible we considered estimates that were broken down by the calendar time of estimation. We additionally wanted to ensure the results weren’t impacted by evaluating different disease outcomes or booster immunogens, and therefore initially only considered vaccine effectiveness estimates against severe disease following XBB.1.5 booster administration. This left us with 27 vaccine effectiveness estimates from 17 studies conducted over a one year period.

As outlined in the methods, we fit a meta-analytic model to log(1-VE), where VE is the reported vaccine effectiveness. This means that a given change in the predictor(s) produces a smaller absolute change in vaccine effectiveness at high effectiveness and a larger change at low effectiveness, consistent with patterns observed in practice. The underlying definition for (1-VE) has some heterogeneity across studies, variously relating to odds ratios, hazard ratios or risk ratios; for simplicity, we refer to it throughout as a relative risk (RR), while noting it may represent an odds or hazard ratio in some cases. In all studies, however, an increase in the modelled effect (referred to as relative risk), corresponds to a decrease in vaccine effectiveness.

We found evidence that the relative risk of severe disease following XBB.1.5 booster administration increased by 4.5% [95%CI 1.7-7.4] per calendar month from mid 2023 to mid 2024 (p=0.0014, Figure 3A, Supplementary Figure 1A and Supplementary Table 1). This increase in relative risk was not significantly different for other disease outcomes (Wald test on interaction terms involving outcome category: χ^2^=0.66, df=2, p=0.72). There were not enough available data on vaccine effectiveness in the first month following JN.1 / KP.2 booster to conclusively show that the relative risk of disease increased with time of study for non-XBB.1.5 boosters (6 observations from 4 studies), however a trend towards an increase in relative risk over time was observed (Figure 3C and Supplementary Table 2). Additionally there was no evidence that the increase in relative risk over time differed for the different booster immunogens (p=0.86 for interaction term involving booster immunogen, Supplementary Table 3).

**Figure 3.**
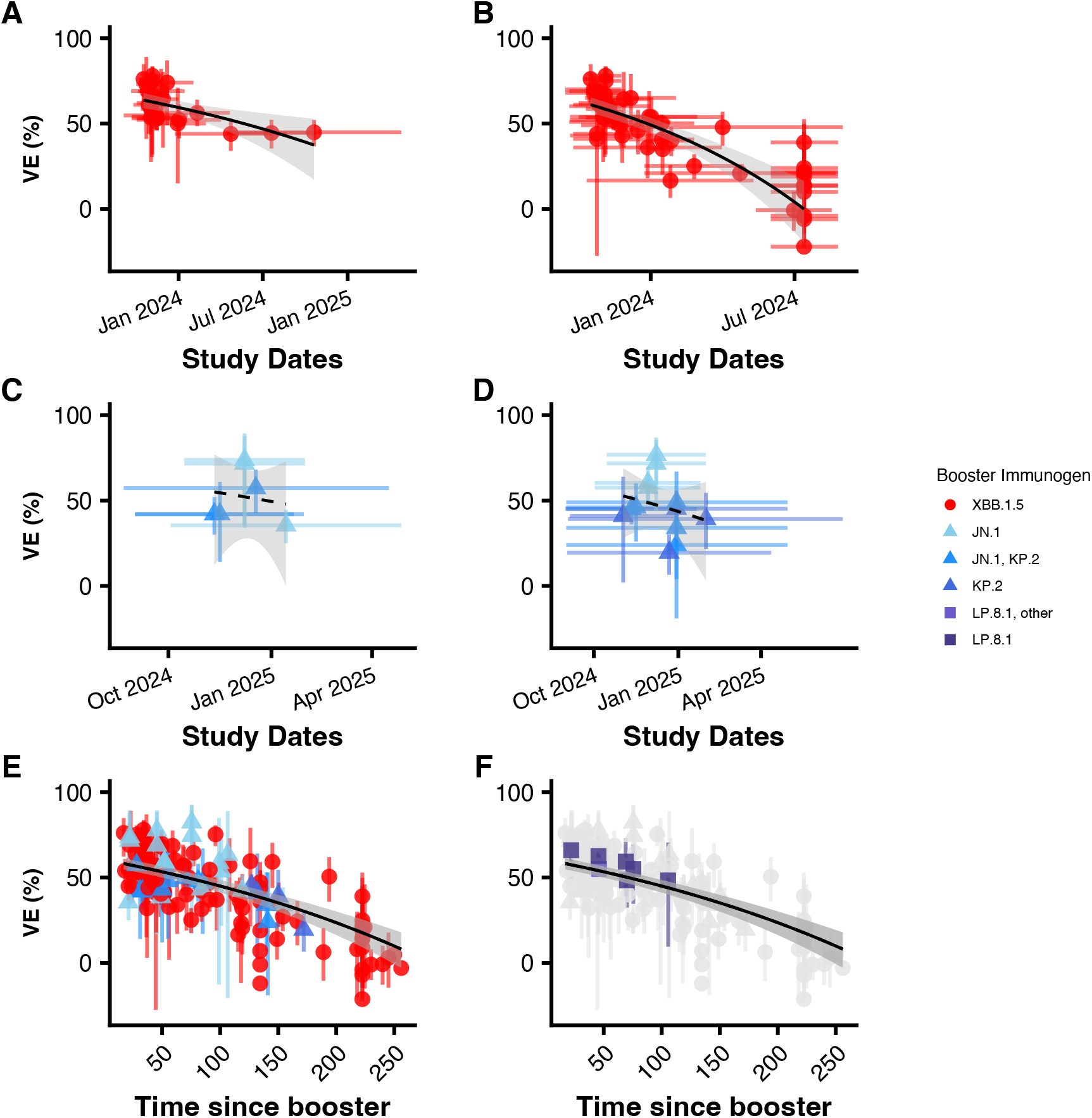
(A-D) Decreasing vaccine effectiveness over calendar date following boosting with the XBB.1.5 immunogen (A and B) and JN.1 or KP.2 immunogens (C and D). Panels A and C show VE (and fits to VE) calculated over the first month following boosting. Panels B and D show VE (and fits to VE) calculated over the entire study duration. Note that some studies report vaccine effectiveness over the first month but not over the entire study duration (and vice versa, so the studies contributing to each panel are not necessarily the same). Horizontal bars show the range of dates over which the study was conducted, with vaccine effectiveness being plotted at the midpoint of the study. (E-F) Decreasing vaccine effectiveness over time since boosting following boosting with the XBB.1.5, JN.1 and KP.2 immunogens (E) and highlighting VE following vaccination with the LP.8.1 immunogen (F). For all panels, vertical error bars show the reported 95% confidence intervals on the vaccine effectiveness estimates.

We note that even when we did not restrict our analysis to the first month following booster administration and instead considered vaccine effectiveness calculated over the entire duration of the study, we found the same result. That is, we found a significant correlation between the relative risk of severe disease following XBB.1.5 boosting and calendar time of the study (19 studies, p<0.0001, Figure 3B, Supplementary Figure 1B and Supplementary Table 4) and a similar (non-significant) trend following boosting with a JN.1 or KP.2 containing vaccine (Figure 3D, Supplementary Figure 1D and Supplementary Table 5).

### What is the combined impact of antibody waning and antigenic drift?

The waning of vaccine effectiveness after booster administration has been reported in a number of previous studies^4,8^. Our dataset allowed a model-based meta-analysis of vaccine protection across all the available data following vaccination with an XBB.1.5, JN.1 or KP.2 containing mRNA vaccine. By considering the reported vaccine effectiveness at different times following booster administration we were able to show that vaccine effectiveness waned significantly over time since booster administration (relative risk of severe disease increased by 10.4% [95%CI 8.6-12.2] per month after booster administration, Figure 3E and Supplementary Table 6), and that the degree of waning did not differ significantly for different disease outcomes (symptomatic disease or death) (joint Wald test on interaction terms between time since booster and disease outcome: χ^2^=5.33, df=2, p=0.07, Supplementary Table 7). For full details see Supplementary Results.

Combined with our findings above that vaccine effectiveness decreases over calendar time, this suggests that both antibody waning and antigenic drift may independently impact vaccine effectiveness. Our aggregated dataset contained estimates of the effectiveness of boosters containing different immunogens and administered during different time periods. We calculated the difference between the calendar time of the study and the date at which the booster immunogen was first reported, and used this as a proxy for antigenic drift. Thus, our measure of antigenic drift became the time that had elapsed between the booster immunogen emergence and the calendar time of the study.

We considered all available data on vaccine effectiveness after boosting with any immunogen (217 estimates from 44 studies). Where possible we used vaccine effectiveness estimates that were broken down by either the time since booster administration or the calendar time of estimation. We set up a mixed effects meta-regression model that included: (i) the time since booster administration; (ii) the time between booster immunogen emergence and the study date; (iii) the outcome against which effectiveness was reported; and (iv) the immunogen contained within the booster. Since 4 of the 13 studies (31%) reporting on effectiveness following a JN.1 or KP.2 containing vaccine combined data from individuals vaccinated with either of these immunogens we grouped these two vaccines together in our analysis.

We found that after accounting for the time since immunogen emergence, the immunogen contained within the booster adds little explanatory power to the model (p=0.11) and therefore removed it from our main analysis (model results including booster immunogen included as a variable are shown in Supplementary Table 8). We will hereafter refer to the model containing the two time-related variables and the disease outcome variable as the “combined” model. We found that both the time since booster administration and the time since booster immunogen emergence were significantly correlated with relative risk (p<0.0001 for both). Model parameters for the combined model are given in Supplementary Table 9.

Using this combined model, we estimated that the relative risk of disease increased by 9% [95%CI 7.4-10.5] per month since booster administration (i.e. due to antibody waning, dashed lines in Figure 4) and by a further 2.4% [95%CI 1.2-3.6] per month since immunogen emergence (i.e. due to antigenic drift of the circulating variant, solid line in Figure 4).

**Figure 4.**
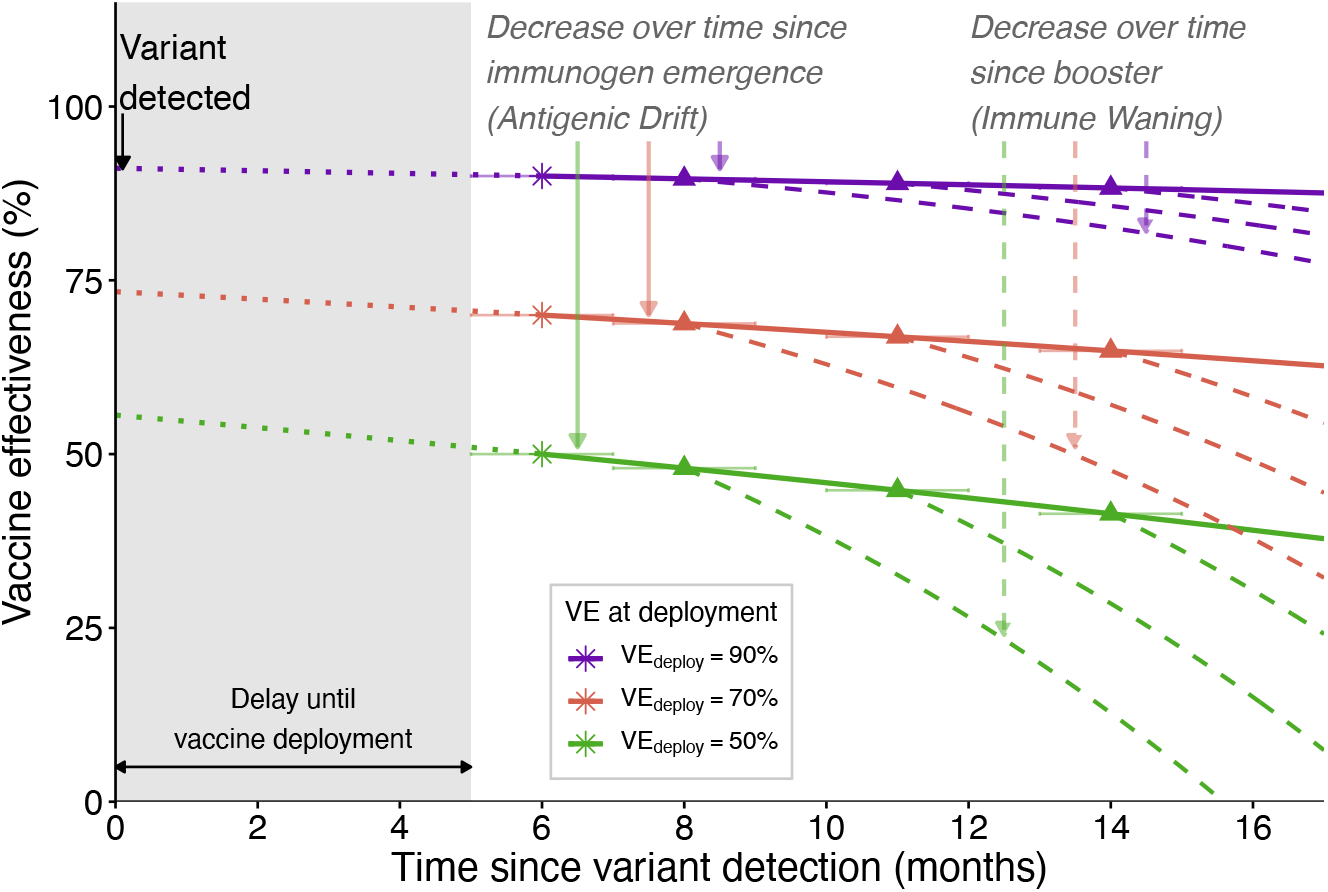
Schematic showing how vaccine effectiveness wanes due to both the time since booster administration (steep dashed lines) and time since variant emergence (shallow solid lines). Grey shaded area represents the time between when the variant was first detected and the time when the booster immunogen containing the variant was first deployed. Stars represent the vaccine effectiveness at the time of booster deployment. Dotted lines in grey area represent the hypothetical vaccine effectiveness if the booster vaccine was deployed earlier. Triangles represent the midpoint of the times over which the hypothetical studies were conducted, horizontal bars represent the duration of the hypothetical studies. Different coloured lines show trajectories for vaccines with different vaccine effectiveness at the time of deployment.

This leads us to conclude that, at the same calendar time, a vaccine containing a more recent immunogen will provide a higher level of protection (since less time will have elapsed since the immunogen’s emergence).

We found no clinical studies that directly compared the effectiveness of two different booster immunogens head-to-head, making it difficult to estimate the benefits of updating booster immunogens. Using the parameters from our analysis we can directly estimate the relative effectiveness of old and new booster immunogens in the same time window. We note that over the past few years new variants (that have eventually been selected as vaccine candidates), have emerged every 10-14 months (e.g. JN.1 and KP.2 were first detected 10 and 14 months after first detection of XBB.1.5, respectively). Therefore, based on the modelling above, we would predict that when administered at the same calendar time, the relative risk associated with receiving an XBB.1.5 vaccine would be 24-33% higher than the relative risk associated with a JN.1 or KP.2 vaccine.

### Agreement with LP.8.1 Vaccine Effectiveness Studies

The ViewHub systematic review concluded in December 2025, and as a result, did not include any studies reporting on vaccine effectiveness following boosting with the LP.8.1 vaccine immunogen. Through ongoing literature review in 2026 we identified 6 studies^9-14^ reporting on LP.8.1 booster effectiveness. We note that 2 of these studies reported vaccine effectiveness in a cohort in which only some of the participants were confirmed to have been given an LP.8.1 containing vaccine ^10,14^. We extracted vaccine effectiveness estimates from these 6 studies (broken down over the time since booster administration where possible) to assess whether they aligned with predictions from our modelling.

It was not possible to evaluate potential declines in effectiveness of the LP.8 booster vaccines over the time since the LP.8.1 immunogen emergence because all identified studies were conducted at very similar times.

However, we found that the effectiveness of the LP.8.1 booster immunogen exactly aligned with the effectiveness predicted from our earlier analysis, and that it waned in almost exactly the same manner as that estimated for XBB.1.5 and JN.1/KP.2 containing vaccines. This can be seen for severe outcomes in Figure 3F and Supplementary Figure 1F (and for other outcomes in Supplementary Figure 2G-I) by the almost exact overlay of the reported effectiveness of the LP.8 boosters over the time since booster administration and our original model.

## Discussion

The continued evolution of SARS-CoV-2 variants has necessitated ongoing updates to the COVID-19 vaccine immunogen. While it is clear that these updates elicit stronger immunological responses to circulating variants^6^, it is less obvious that successive updates to the vaccine antigen composition translate into meaningful improvements in vaccine effectiveness, beyond the benefits conferred by boosting alone^15^. Using a meta-analysis of all vaccine effectiveness studies identified by the ViewHub systematic review^7^, we have shown that both antibody waning (represented by the time since vaccine administration) and antigenic drift (represented by the time since booster immunogen emergence) independently impact vaccine effectiveness.

We identified that vaccine effectiveness wanes as the time since vaccine administration increases (corresponding to an increase in 1-VE of 9% per month), which is likely to be the result of antibody waning^4,16,17^. In parallel, there is an independent reduction in vaccine effectiveness as the time between the first emergence of the variant increases (RR increase of 2.5% per month), which likely reflects progressive antigenic drift and thus a reduced immunological cross-reactivity to the newly circulating variant.

Together, these findings indicate that sustained vaccine effectiveness depends on both the durability of vaccine-induced immunity and continued antigenic updates of vaccine immunogens to continue to improve antibody responses to evolving variants.

The fact that both the time since vaccination and the time at which a study is performed can impact estimates of vaccine effectiveness means that simple comparisons of effectiveness may not capture the true benefit of vaccine updates. An ideal study to compare vaccine effectiveness of different immunogens would administer each of these vaccines to different cohorts of people at the same time and would compare the vaccine effectiveness of the different products. This would ideally be a placebo controlled double blinded study, although a study run at the same chronological time would suffice. In practice, however, no such studies have been implemented. Indeed, studies using different booster immunogens were almost never run at similar times, likely reflecting the fact that boosters containing each immunogen were only available for use over specific time periods.

This work has a number of limitations. Firstly, we note that the vaccine effectiveness estimates used were the adjusted published estimates and were not calculated directly from individual study event counts and demographic information. These estimates were calculated in different ways, variously depending on calculations of odds, hazard and risk ratios. This approach enabled us to include a larger body of evidence and allowed us to rely on adjustments made by study investigators (who are more aware of any study-specific confounders). Direct analysis of original event data would have allowed for more accurate model fitting, and may have reduced model associated error, however this information was not available in ViewHub, nor was it reported for all studies.

Secondly, we note that there are substantially fewer studies reporting on vaccine effectiveness of JN.1 and KP.2 boosters than of XBB.1.5 boosters, and fewer still reporting on vaccine effectiveness of LP.8.1 vaccines. Although all published data consistently supported the conclusion that vaccine effectiveness against disease and death decreases over both the time since booster administration and the time since the booster immunogen emergence, incorporation of additional studies of newer vaccine formulations would improve the certainty of our estimates. We note however that vaccine effectiveness estimates from recently published LP.8.1 studies closely matched model predictions despite not being included in the original model fitting, providing independent validation of our model.

Finally, our analysis did not explicitly account for changes in population immunity arising from repeated infection and prior vaccinations as exposure / vaccination history was not consistently reported in the included studies. Since such immunity has increased over time, the incremental benefit measured in later vaccine effectiveness studies may underestimate the protective effect that would have been observed at earlier stages of the pandemic.

Here we have presented a comprehensive meta-analysis of COVID-19 booster vaccine effectiveness for boosters containing XBB.1.5, JN.1 and KP.2 immunogens. We have shown that both antibody waning and antigenic drift contribute to declines in vaccine effectiveness, and that updating vaccines to provide a closer match between the vaccine immunogen and circulating variant is likely to lead to improvements in vaccine effectiveness.

## Methods

### Study identification

Studies of COVID-19 booster vaccine effectiveness were identified by using the VIEW-hub living systematic review of COVID-19 Vaccine Effectiveness Studies (IVAC, 2026). The dataset was maintained by the International Vaccine Access Center and the Bloomberg School of Public Health at Johns Hopkins University until 31 December 2025. In addition, we compiled COVID-19 vaccine effectiveness (VE) studies reported in the preprint literature, peer-reviewed publications, and official reports from the start of the pandemic up until 31 December 2025.

#### Inclusion Criteria

Studies were included in our meta-analysis if they reported vaccine effectiveness in a healthy population following vaccination with an XBB.1.5, JN.1, KP.2 or LP.8 containing vaccine.

#### Data extraction

Full-text articles of all eligible studies were accessed, and data were checked (by AM) for consistency with the data that was provided for download from ViewHub. In addition to the variables available in ViewHub’s downloadable spreadsheet, we extracted information, where reported, on booster immunogen, the range in the time since booster vaccination (either interquartile range (IQR) or entire range), mean or median cohort age, study period (start and end dates), and the period over which effectiveness was assessed (start and end dates corresponding to participant recruitment or the observation period used in the analysis, if different from the study period). All additional data was extracted and checked by AM and WJ.

In some instances we used ChatGPT^18^ for simple calculations or to locate relevant data within publications. All such calculations and data were independently manually verified by AM. When data were only presented in a figure such data were extracted from the figures using WebPlotDigitizer^19^ (version 4.8).

#### Vaccine Effectiveness

Vaccine effectiveness (VE) estimates were extracted for both the entire study duration (overall estimate) and stratified by time since booster administration (stratified-time) and the circulating variant (stratified-variant) where reported. Vaccine effectiveness estimates for immunocompromised individuals, high-risk populations, or pregnant individuals were excluded, as were estimates measured within 14 days of booster administration.

“Overall” vaccine effectiveness estimates generally had smaller confidence intervals, as they were based on more cases and assessed over a longer time frame. In contrast, stratified estimates provided greater resolution when assessing waning vaccine effectiveness (over time since booster administration) and changes in vaccine effectiveness over calendar time (i.e. when there were different variants circulating).

Unless otherwise stated, analysis was performed on vaccine estimates stratified by time since booster administration (stratified-time) and by circulating variant (stratified-variant). Results of fitting to non-stratified vaccine effectiveness estimates are presented in the supplementary materials.

In some analyses we were interested in vaccine effectiveness estimates close to the time of booster administration (and therefore prior to any significant waning having occurred). In this case, we considered only VE estimates closest to one month after booster administration. We determined this to be the estimate taken closest to 30 days after booster administration, with a median (or mean) time since booster administration of no less than 14 days and no more than 45 days, and with the upper bound of the reported time since booster administration required to be under 60 days.

Where multiple VE estimates were reported for different population subgroups within a study, we identified a primary analysis population corresponding to the principal population analysed by the study authors. Unless otherwise specified, only estimates from this primary population were included in the main analysis to avoid multiple correlated observations from the same study. For the sensitivity analysis restricted to adult populations, vaccine effectiveness estimates from studies with a primary population that included both adults and children were instead replaced by an adult-only vaccine effectiveness estimate (where available).

#### Determining the date of emergence for a booster immunogen

Since the observed date of a variant’s emergence depends on both the intensity of sampling and sequencing, and the propensity for sampled sequences to be submitted to public databases, the “true” emergence date of a given variant cannot be directly determined. The pango-designation GitHub repository^20^ is the authoritative, version-controlled record of Pango lineage designations, in which accepted lineages are recorded. We therefore used the date on which each corresponding Pango lineage^21^ was first added to this repository as a proxy for its emergence date (Table 1). These designation dates provide reliable upper bound on emergence because, by definition, a lineage must have existed before it could be identified and designated.

**Table 1.** Designation (classification) dates for booster immunogens considered in this work (Pango lineages: XBB.1.5, JN.1, KP.2, LP.8.1).

| Pango Lineage | Pango designation date |
| --- | --- |
| XBB.1.5 | 8 November 2022 |
| JN.1 | 29 September 2023 |
| KP.2 | 4 March 2024 |
| LP.8.1 | 8 November 2024 |

#### Calendar time of study and time since immunogen emergence

The point estimate for the calendar time of a study was the midpoint of the dates over which the VE was assessed in the study.

The time since immunogen emergence was calculated as the time that had elapsed between the date of emergence for that booster immunogen (see Table 1) and the point estimate for the calendar time of the study.

### Statistical analysis

#### Meta-Analytic Regression Models

We analysed the impact of different variables on vaccine effectiveness using meta-analytic mixed-effects models implemented using the *rma*.*mv* function in the *metafor* package in R (v4.4.2; R Core Team 2021).

We used a hierarchical mixed-effects model fitted using restricted maximum likelihood (REML). We set the sampling variance of the outcome measure to be the square of the standard error of the outcome measure. The standard error of the outcome measure,

*SE*, was determined as *SE* = (*M*_*u*_ − *M*_*l*_)/(2×1.96) where *M*_*u*_ and *M*_*l*_ are the upper and lower 95% confidence intervals for the outcome measure, respectively. The model intercept was allowed to vary at both the study level (to account for heterogeneity in the underlying effect between studies) and at the estimate-within-study level (since some studies contributed multiple estimates, e.g. for different age groups or countries). Data with missing information for variables required in a given analysis were excluded from that analysis. Fixed effects considered for inclusion are specified in the relevant section of the manuscript.

Unless otherwise specified, the outcome variable was ln(1 − *VE*) (i.e. the natural logarithm of 1 − *VE*, where VE is the vaccine effectiveness). Analyses using vaccine effectiveness itself as the outcome variable are reported in the supplementary materials.

Comparisons of vaccine effectiveness using different booster immunogens were also performed using meta-analytic mixed-effects models in which booster immunogen was included as the sole predictor variable.

Joint Wald tests were used to assess the statistical significance of included variables and interaction terms, with p-values <0.05 considered statistically significant.

## Data Availability

All data and code will be made available after publication

## Notes

### Competing Interest Statement

The authors have declared no competing interest.

### Author Declarations

The study used ONLY openly available human data that were originally located at ViewHub (https://view-hub.org/)

