## Supplementary materials for "Separating the effects of immune waning and viral evolution on COVID-19 vaccine effectiveness reveals improved effectiveness of updated vaccines"

### 1. Supplementary Results

#### *1.1. Do we see evidence that vaccine effectiveness wanes over time since boosting?*

It has previously been shown that vaccine effectiveness wanes over time since booster administration. We considered the reported vaccine effectiveness at different times following booster administration in the studies for which this was reported, or over the duration of the study if effectiveness was not broken down by time since booster administration.

Since vaccine effectiveness varies against different outcomes, we initially analysed effectiveness reported against severe outcomes only, and found that across all identified data, the relative risk of severe disease increased by 10.4% [95%CI 8.6-12.2] per month after booster administration. The resulting prediction of vaccine effectiveness over time since booster administration is shown in Figure 3E of the main manuscript, while a figure showing the relationship between relative risk and time since booster administration is given in Supplementary Figure 1E. Full model parameters are presented in Supplementary Table 6.

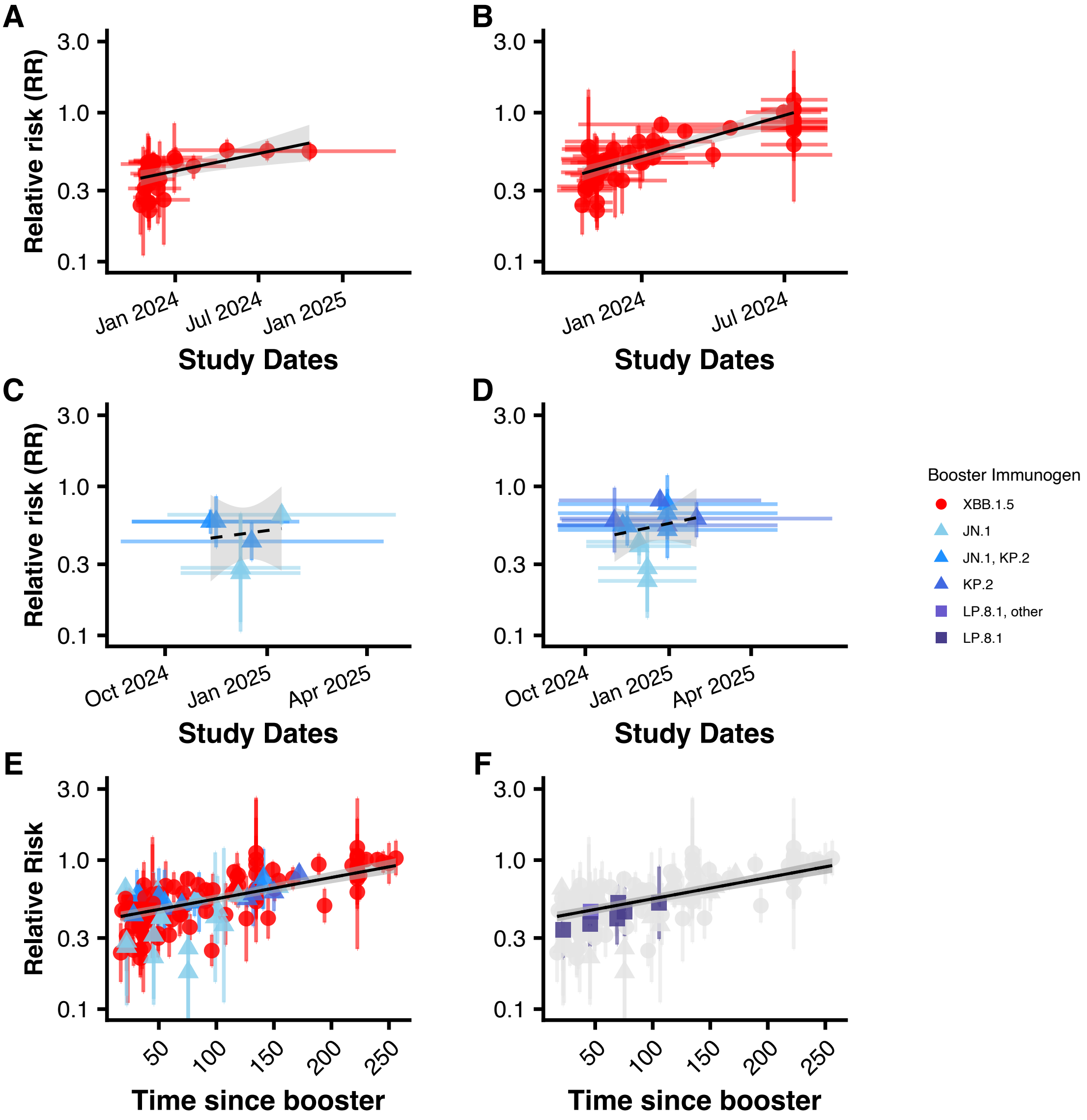


*Supplementary Figure 1 (A-D) Increasing relative risk over calendar date following boosting with the XBB.1.5 immunogen (A and B) and JN.1 or KP.2 immunogens (C and D). Panels A and C show RR (and fits to RR) calculated over the first month following boosting. Panels B and D show RR (and fits to RR) calculated over the entire study duration. Note that some studies report vaccine effectiveness over the first month but not over the entire study duration (and visa-versa, so the studies contributing to each panel are not necessarily the same). Horizontal bars show the range of dates over which the study was conducted, with vaccine effectiveness being plotted at the midpoint of the study. (E-F) Increasing relative risk over time since boosting following boosting with the XBB.1.5, JN.1 and KP.2 immunogens (E) and highlighting RR following vaccination with the LP.8.1 immunogen (F). For all panels, vertical error bars show the reported 95% confidence intervals on the relative risk estimates.*

We additionally tested whether the association between relative risk and time since booster administration differed by the disease outcome (symptomatic disease or death) and found weak evidence that it did (joint Wald test on interaction terms between time since booster and disease outcome: $\chi^2$=5.33, df=2, p=0.07). There was no evidence of a difference in symptomatic compared to severe disease, however we found evidence that the relative risk of death increased more rapidly, increasing by 4.8% [95%CI 0.7-9] more per month since booster administration compared to severe disease. A figure showing the results of this model is provided in Supplementary Figure 2 along with estimated parameters in Supplementary Table 7.

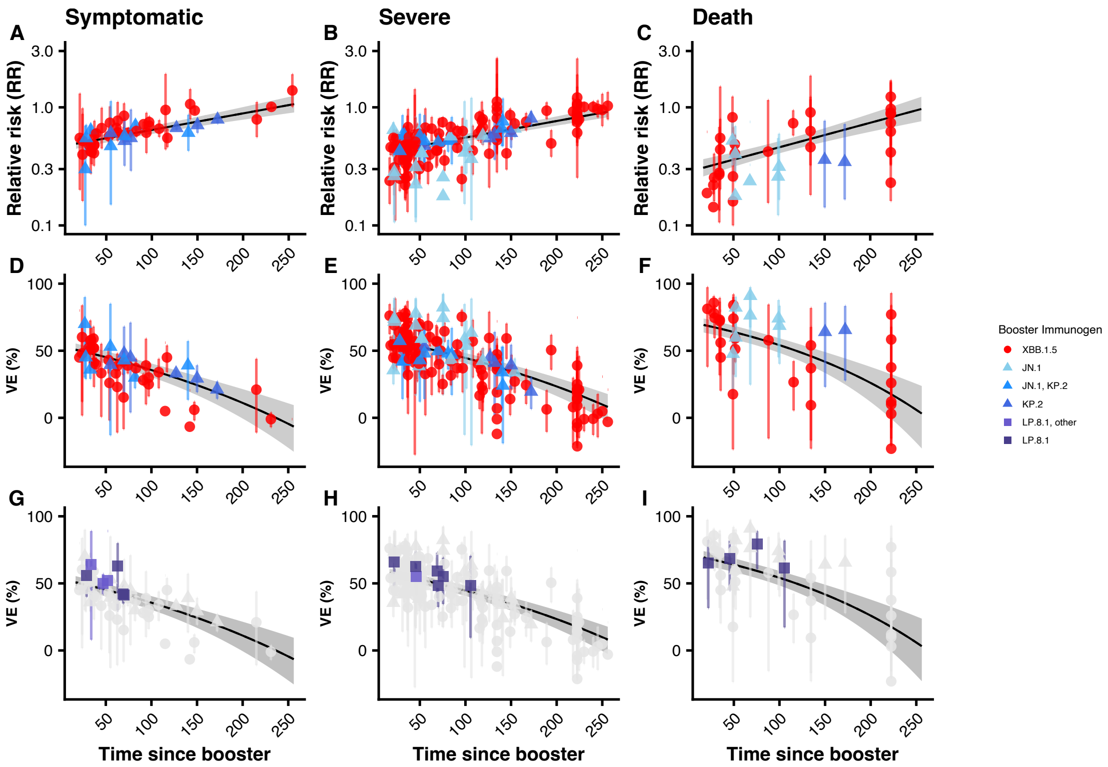


*Supplementary Figure 2 Plots showing the increase in relative risk (panels A-C) and decrease in vaccine effectiveness (panels D-F) over time since boosting, for symptomatic disease (panels A and D) severe disease (panels B and E) and death (panels C and F). Lines show the result of a meta-analytic linear regression model fit to the natural log of the relative risk. Dependant variables are the time since booster administration, outcome, and the interaction between outcome and the time since booster administration. Panels G-I show the same plots and model fit as panels D-F, overlaid with the vaccine effectiveness for LP.8.1 containing booster immunogens.*

#### *1.2. Results of combined model that includes a variable for booster immunogen*

In this section we consider the results of including a variable for the booster immunogen in our combined model. This model, which we will refer to as the "booster-combined" model, containing the two time-related variables, the immunogen contained within the booster and the outcome variable. We found that both the time since booster administration and the time since booster immunogen emergence were significantly correlated with relative risk ($p<0.0001$ for both). Model parameters for the "booster-combined" model are given in Supplementary Table 8.

Using this booster-combined model, we estimated that the relative risk of disease increased by 8.6% [95%CI 7-10.2] per month since booster administration (i.e. due to antibody waning) and by a further 3.1% [95%CI 1.6-4.7] per month since immunogen emergence (i.e. due to antigenic drift of the circulating variant).

#### ***1.3. Results of fitting to vaccine effectiveness directly***

When fitting our model to the vaccine effectiveness directly (rather than to the log of the relative risk), we found the results below:

##### *1.3.1. Do we see evidence that vaccine effectiveness wanes over time since boosting?*

Across all identified data, the vaccine effectiveness against severe disease decreased by 6.3% [95%CI 5.3-7.3] per month after booster administration (Supplementary Figure 3A and D).

We additionally tested whether the association between vaccine effectiveness and time since booster administration differed by the disease outcome (symptomatic disease or death) and found no evidence that it did (joint Wald test on interaction terms between time since booster and disease outcome: $\chi^2=0.66$, df=2, p=0.72).

##### *1.3.2. Is vaccine effectiveness impacted by the time at which a study is conducted?*

We first considered the relationship between vaccine effectiveness estimates over the first month following booster administration and the time when a study was conducted. We found evidence that the vaccine effectiveness against severe disease following XBB.1.5 booster administration decreased by 2% [95%CI 0.9-3.2] per calendar month from mid 2023 to mid 2024 (p=3e-04, Supplementary Figure 3B and E).

This decrease in vaccine effectiveness was not significantly different for other disease outcomes (Wald test on interaction terms involving outcome category: $\chi^2=0.34$, df=2, p=0.84).

We note that even when we did not restrict our analysis to the first month following booster administration and instead considered the effectiveness aggregated over the entire duration of the study, we found the same result. That is, we found a significant correlation between the vaccine effectiveness against severe disease following XBB.1.5 boosting and calendar time of the study (19 studies, p<0.0001). , Supplementary Figure 3C and F)

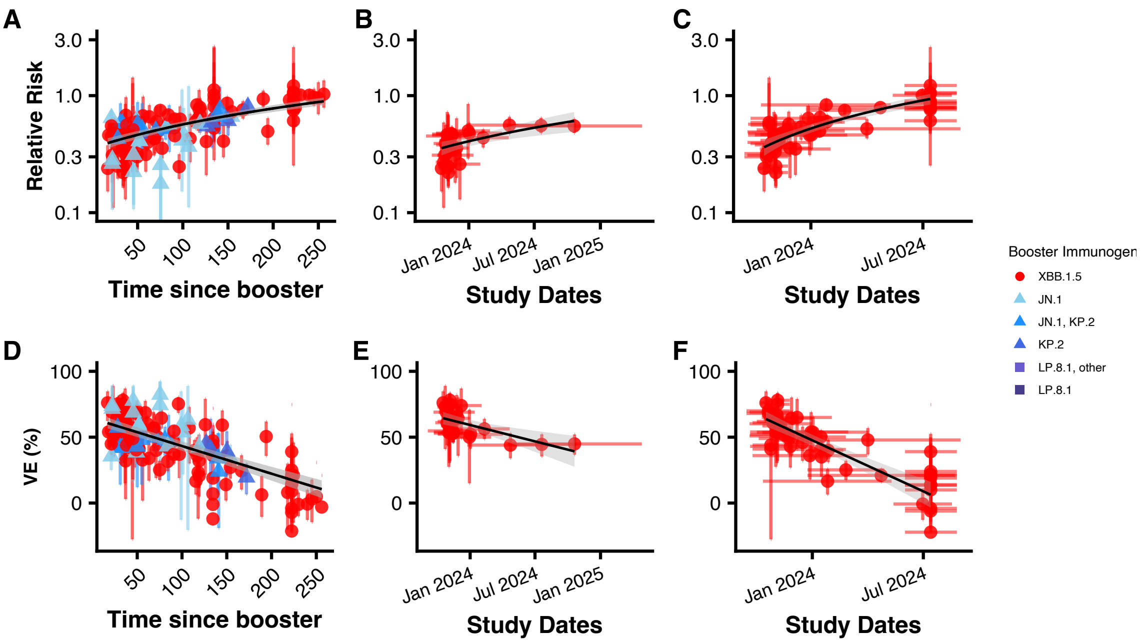


*Supplementary Figure 3 Plots showing increase in relative risk (panels A-C) and decrease in vaccine effectiveness against severe disease (panels D-F) over (A and D) time since boosting, and (B,C,E,F) calendar date. Panels A and D show results of fitting the linear meta-analytic mixed-effects model to vaccine effectiveness estimates following vaccination with all booster immunogens. Panels B, C, E and F show results of fitting the linear meta-analytic mixed-effects model to vaccine effectiveness estimates obtained following vaccination with the XBB.1.5 booster immunogen only. Panels B and E show fits to vaccine effectiveness estimates calculated within one month of booster administration, Panels C and F show fits to vaccine effectiveness estimates calculated over the entire study duration.*

#### *1.3.3. What is the combined impact of antibody waning and antigenic drift?*

We found that the immunogen contained within the booster was not a significant predictor ($p=0.19$), and therefore we removed it from the model. We then found that both the time since booster administration and the time since booster immunogen emergence were significantly correlated with vaccine effectiveness ($p<0.0001$ for both).

Using this combined model, we estimated that the vaccine effectiveness decreased by 5.6% [95%CI 4.8-6.4] per month since booster administration (i.e. due to antibody waning) and by a further 1.6% [95%CI 0.9-2.2] per month since immunogen emergence.

Therefore, based on the modelling above, we would predict that when administered at the same calendar time, the vaccine effectiveness associated with receiving an XBB.1.5 vaccine would be 16-23% lower than the vaccine effectiveness associated with a JN.1 or KP.2 vaccine.

#### *1.4. Analyses using the non-stratified vaccine effectiveness estimates*

When considering the reported vaccine effectiveness estimates that were not stratified by circulating variant, we found that across all identified data, the relative risk of severe disease increased by 9.5% [95%CI 7.6-11.4] per month after booster administration. There was alsosome evidence of an association between relative risk and time since booster administration differing by disease outcome (symptomatic or severe disease or death) (joint Wald test on interaction terms between time since booster and disease outcome: $\chi^2$=6.98, df=2, p=0.031). There was no evidence of a difference in symptomatic compared to severe disease, however we found evidence that the relative risk of death increased more rapidly, increasing by 5.3% [95%CI 1.3-9.4] more per month since booster administration compared to severe disease.

Vaccine effectiveness following XBB or JN.1/KP.2 vaccination was not significantly different either over the entire study (Supplementary Figure 4A) or over the first month following booster administration (p=0.9 and p=0.17 respectively).

Over the period from 1st June 2024 to 31st January 2025, effectiveness was significantly higher in cohorts boosted with a JN.1 or KP.2 containing booster compared to those receiving an XBB.1.5-containing vaccine (median effectiveness against severe disease was 57.5% vs 14%; p=0.0063, Supplementary Figure 4C).

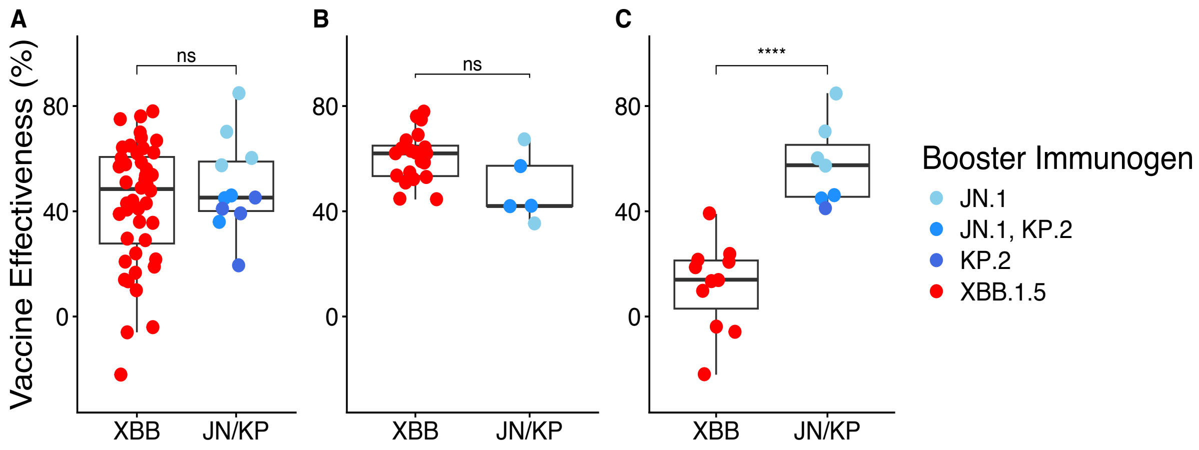


*Supplementary Figure 4 XBB vs JN/KP booster effectiveness evaluated over all variants within a study either (A) over the entire study period in all studies (B) over the first month since booster administration in all studies or (C) over the entire study period in studies conducted wholly between 1st June 2024 and 31st January 2025.*

#### *1.5. Sensitivity analyses*

##### *1.5.1. Sensitivity analysis 1 – Only including estimates soon after booster administration.*

In this analysis, we applied the combined to vaccine effectiveness estimates that were estimated as close to one month after booster administration as possible (and no more than two months after booster administration). In this way, we hoped to avoid any potential interaction between chronological time and time since booster administration. As expected, the time since booster administration was no longer a significant predictor of vaccine effectiveness in this dataset (p=0.73), Supplementary Table 10). Other variables (time since booster immunogen emergence and disease outcome) remained significant predictors in the model when removing the time since booster administration, with a similar estimate being obtained for the change in relative risk over the time since booster immunogen emergence when compared to the combined model from the main analysis (2.4% [95%CI 1.2-3.6] per month in the main model vs 3.1% [95%CI 0.7-5.6] in the restricted model applied to this dataset of early vaccine effectiveness estimates, Supplementary Figure 5 and Supplementary Table 11).

#### *1.5.2. Sensitivity analysis 2 – Including outcome as an interaction term with time related variables.*

It is possible that using one model for all three outcome categories (symptomatic disease, severe disease and death) is an oversimplification and that the outcome category against which vaccine effectiveness is evaluated not only impacts the overall relative risk of disease but also affects the relationship between this risk and the time since booster administration or time since variant emergence. We therefore also considered models that included interaction terms between these variables and the outcome category.

We considered interactions between disease outcome and the time-related variables of the combined model (Supplementary Table 12). We found no evidence that the change in vaccine effectiveness over time differed for symptomatic compared to severe disease (joint Wald test on interactions terms incorporating symptomatic disease: $\chi^2$=0.26, df=2, p=0.878). However, we found evidence of a greater decrease in the vaccine effectiveness over time the outcome of death (joint Wald test on interactions terms incorporating death :$\chi^2$=11.54, df=2, p=0.0031). This may represent a similar decrease in vaccine effectiveness against all disease outcomes, however since the effectiveness against death starts higher, this translates to a greater decrease in relative risk.

#### *1.5.3. Sensitivity analysis 3 – including age*

It is often assumed that vaccine effectiveness may be lower in older age groups. We therefore tested whether age was a significant factor affecting our conclusions regarding the relative risk of disease at different times or following vaccination with different booster immunogens. We repeated our analysis, this time including the median age in the population as a variable in the model. Although the vaccine effectiveness was predicted to increase with increasing age, this did not reach statistical significance (p=0.38). In addition, it did not dramatically impact the other parameters of the model. In addition, it did not dramatically impact the other parameters of the model (Supplementary Table 13).

We also re-fit the combined model, restricting the dataset to include only estimates of vaccine effectiveness in cohorts that did not include participants under 18 years of age. We found the results were almost identical to those from our main analysis and there was a less than -101% difference the estimated change in relative risk over the time since booster administration or immunogen emergence when fitting to this adult-only dataset (Supplementary Table 14).

*1.5.4. Sensitivity analysis 4 – including location of study or study type*

We next wanted to determine that our conclusions were not impacted by the location where the study was conducted, or by the type of study that was conducted. We therefore re-ran our combined model including a variable for either the location of the study (Europe, North America or Asia Pacific) or the study type (test-negative case-control study or cohort study).

We found that neither the study location nor the study type were significant predictors in our model (joint Wald test on terms related to study location in a model including study location: $\chi^2$=5.43, df=2, p=0.066 and joint Wald test on terms related to study type in a model including study type: $\chi^2$=0.38, df=1, p=0.54), Supplementary Tables 15 and 16).

*1.5.5. Sensitivity analysis 5 – considering JN.1 and KP.2 boosters separately*

In the main analysis we considered both JN.1 and KP.2 boosters together as 4/13 studies using these booster immunogens were of a cohort that could have been boosted with either immunogen. However, we identified 9 studies that reported vaccine effectiveness separately for JN.1 or KP.2 containing boosters.

We therefore next applied our combined model to these 9 studies that reported on vaccine effectiveness following boosting with only a single booster immunogen (i.e. XBB1.5, JN.1 or KP.2) and included a variable for booster immunogen in the model (Supplementary Table 17). Once again, we did not find any evidence that booster immunogen was a significant predictor of vaccine effectiveness (joint Wald test on terms related to study booster immunogen: $\chi^2$=1.72, df=2, p=0.42), confirming our previous finding that there is no evidence of an association between relative risk and booster immunogen.

### 2. Other Supplementary Figures

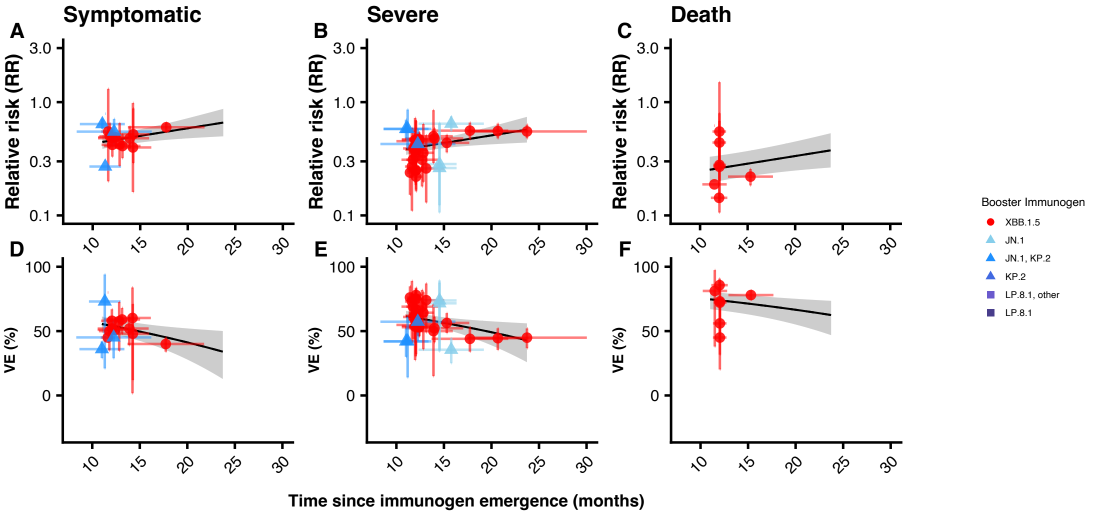


*Supplementary Figure 5* *Plots showing the increase in relative risk (panels A - C) and decrease in vaccine effectiveness (panel D - F) over time since booster immunogen emergence, for symptomatic disease (panels A and D) severe disease (panels B and E) and death (panels C and F). All the panels show RR and VE calculated closest to one month of booster administration. Lines show the result of a meta-analytic linear regression model fit to the natural log of the relative risk. Dependant variables are the time since booster immunogen emergence and outcome.*

### 3. Supplementary Tables

| Parameter | Estimate (95% CI) | p-value |
|---|---|---|
| Intercept | 0.2372 (0.1677, 0.3354) | <0.0001 |
| Time (months) | 1.045 (1.017, 1.074) | 0.0014 |

*Supplementary Table 1 Results of meta-analytic model fitted to severe vaccine effectiveness data in the first month following XBB.1.5 vaccination. Data includes vaccine effectiveness estimates closest to one month after booster administration, stratified by circulating variant (total number of data points n = 27). Baseline values correspond to severe disease. Model specification: ln(1-VE) ~ calendar time.*

| Parameter | Estimate (95% CI) | p-value |
|---|---|---|
| Intercept | 0.09370 (0.000, 11400) | 0.6919 |
| Time (months) | 1.072 (0.6548, 1.754) | 0.7832 |

*Supplementary Table 2 Results of meta-analytic model fitted to severe vaccine effectiveness data in the first month following vaccination with the JN.1 or KP.2 booster immunogen. Data includes vaccine effectiveness estimates closest to one month after booster administration, stratified by circulating variant (n = 6). Baseline values correspond to severe disease. Model specification: ln(1-VE) ~ calendar time.*

| Parameter | Estimate (95% CI) | p-value |
|---|---|---|
| Intercept | 0.2427 (0.1698, 0.3470) | <0.0001 |
| Time (months) | 1.043 (1.014, 1.072) | 0.0033 |
| JN.1 or KP.2 immunogen | 0.4694 (0.001100, 195.2) | 0.8058 |
| Interaction Time:Immunogen | 1.023 (0.7942, 1.317) | 0.8612 |

*Supplementary Table 3 Results of meta-analytic model fitted to severe vaccine effectiveness data in the first month following vaccination with any immunogen. Data includes vaccine effectiveness estimates closest to one month after booster administration, stratified by* booster immunogen *(n = 33). Baseline values correspond to XBB.1.5 booster immunogen. Model specification: ln(1-VE) ~ calendar time * booster immunogen.*

| Parameter | Estimate (95% CI) | p-value |
|---|---|---|
| Intercept | 0.1420 (0.1012, 0.1991) | <0.0001 |
| Time (months) | 1.110 (1.082, 1.140) | <0.0001 |

*Supplementary Table 4 Results of meta-analytic model fitted to severe vaccine effectiveness data over the entire study period following vaccination with the XBB.1.5 booster immunogen. Data includes vaccine effectiveness estimates stratified by circulating variant (n = 48). Model specification: ln(1-VE) ~ calendar time.*

| Parameter | Estimate (95% CI) | p-value |
|---|---|---|
| Intercept | 0.06610 (0.0002000, 25.90) | 0.3725 |
| Time (months) | 1.092 (0.8497, 1.403) | 0.4917 |

*Supplementary Table 5 Results of meta-analytic model fitted to severe vaccine effectiveness data over the entire study period following vaccination with the JN.1 or KP.2 booster immunogen. Data includes vaccine effectiveness estimates stratified by circulating variant (n = 13). Model specification: ln(1-VE) ~ calendar time.*

| Parameter | Estimate (95% CI) | p-value |
|---|---|---|
| Intercept | 0.3958 (0.3649, 0.4294) | <0.0001 |
| Time since booster administration (months) | 1.104 (1.086, 1.122) | <0.0001 |

*Supplementary Table 6 Results of meta-analytic model fitted to severe vaccine effectiveness data only. Data includes vaccine effectiveness estimates stratified by time since booster administration and circulating variant (n = 135). Model specification: ln(1-VE) ~ time since booster.*

| Parameter | Estimate (95% CI) | p-value |
|---|---|---|
| Intercept | 0.4005 (0.3694, 0.4343) | <0.0001 |
| Time since booster administration (months) | 1.102 (1.085, 1.119) | <0.0001 |
| Outcome=Symptomatic | 1.164 (1.047, 1.294) | 0.0050 |
| Outcome=Death | 0.7085 (0.5918, 0.8482) | 0.0002 |
| Interaction<br>Time since booster:Symptomatic | 0.9995 (0.9715, 1.028) | 0.9731 |
| Interaction<br>Time since booster:Death | 1.048 (1.007, 1.090) | 0.0212 |

*Supplementary Table 7 Results of meta-analytic model fitted to all vaccine effectiveness data. Data includes vaccine effectiveness estimates stratified by time since booster administration and circulating variant (n = 217). Baseline values correspond to severe disease. Model specification: ln(1-VE) ~ time since booster * outcome.*

| Parameter | Estimate (95% CI) | p-value |
|---|---|---|
| Intercept | 0.2616 (0.2092, 0.3270) | <0.0001 |
| Time since booster administration (months) | 1.086 (1.070, 1.102) | <0.0001 |
| Time since immunogen emergence (months) | 1.031 (1.016, 1.047) | <0.0001 |
| JN.1 or KP.2 immunogen | 1.111 (0.9756, 1.266) | 0.1123 |
| Outcome=Symptomatic | 1.171 (1.102, 1.245) | <0.0001 |
| Outcome=Death | 0.8274 (0.7465, 0.9171) | 0.0003 |

*Supplementary Table 8 Results of meta-analytic model fitted to vaccine effectiveness data following vaccination with any immunogen. Data includes vaccine effectiveness estimates stratified by time since booster administration and circulating variant (n = 217). Baseline values correspond to severe disease after having received the XBB.1.5 booster immunogen. Model specification: ln(1-VE) ~ time since booster + time since immunogen emergence + booster immunogen + outcome.*

| Parameter | Estimate (95% CI) | p-value |
|---|---|---|
| Intercept | 0.2950 (0.2498, 0.3484) | <0.0001 |
| Time since booster administration (months) | 1.090 (1.074, 1.106) | <0.0001 |
| Time since immunogen emergence (months) | 1.024 (1.012, 1.036) | <0.0001 |
| Outcome=Symptomatic | 1.172 (1.102, 1.247) | <0.0001 |
| Outcome=Death | 0.8298 (0.7480, 0.9205) | 4e-04 |

*Supplementary Table 9 Results of combined meta-analytic model fitted to vaccine effectiveness data following vaccination with any immunogen. Data includes vaccine effectiveness estimates stratified by time since booster administration and circulating variant (n = 217). Baseline values correspond to severe disease. Model specification: ln(1-VE) ~ time since booster + time since immunogen emergence + outcome.*

| Parameter | Estimate (95% CI) | p-value |
|---|---|---|
| Intercept | 0.2545 (0.1490, 0.4347) | <0.0001 |
| Time since booster administration (months) | 1.079 (0.7009, 1.661) | 0.7300 |
| Time since immunogen emergence (months) | 1.032 (1.007, 1.057) | 0.0126 |
| Outcome=Symptomatic | 1.148 (0.9992, 1.319) | 0.0513 |
| Outcome=Death | 0.6607 (0.5136, 0.8499) | 0.0013 |

*Supplementary Table 10 Results of combined meta-analytic model fitted to vaccine effectiveness data in the first month following vaccination with any immunogen. Data includes vaccine effectiveness estimates closest to one month after booster administration, stratified by circulating variant (n = 54). Baseline values correspond to severe disease. Model specification: ln(1-VE) ~ time since booster + time since immunogen emergence + outcome.*

| Parameter | Estimate (95% CI) | p-value |
|---|---|---|
| Intercept | 0.2740 (0.1952, 0.3845) | <0.0001 |
| Time since immunogen emergence (months) | 1.031 (1.007, 1.056) | 0.0109 |
| Outcome=Symptomatic | 1.155 (1.012, 1.319) | 0.0324 |
| Outcome=Death | 0.6570 (0.5122, 0.8428) | 0.0009 |

*Supplementary Table 11 Results of combined meta-analytic model fitted to vaccine effectiveness data in the first month following vaccination with any immunogen. Meta-regression analysis restricted to estimates for symptomatic infection, severe disease, and death outcomes. Data includes vaccine effectiveness estimates closest to one month after booster administration, stratified by circulating variant (n = 54). Baseline values correspond to severe disease. Model specification: ln(1-VE) ~ time since immunogen emergence + outcome.*

| Parameter | Estimate (95% CI) | p-value |
|---|---|---|
| Intercept | 0.3024 (0.2536, 0.3607) | <0.0001 |
| Time since booster administration (months) | 1.091 (1.073, 1.108) | <0.0001 |
| Time since immunogen emergence (months) | 1.022 (1.009, 1.035) | 0.0007 |
| Outcome=Symptomatic | 1.255 (0.9544, 1.650) | 0.1040 |
| Outcome=Death | 0.4253 (0.2643, 0.6845) | 0.0004 |
| Interaction<br>Time since booster:Symptomatic | 1.000 (0.9712, 1.030) | 0.9829 |
| Interaction<br>Time since booster:Death | 1.020 (0.9707, 1.071) | 0.4393 |
| Interaction<br>Time since emergence:Symptomatic | 0.9948 (0.9735, 1.016) | 0.6352 |
| Interaction<br>Time since emergence:Death | 1.037 (1.002, 1.073) | 0.0353 |

*Supplementary Table 12 Results of combined meta-analytic model fitted to vaccine effectiveness data following vaccination with any immunogen. Data includes vaccine effectiveness estimates stratified by time since booster administration and circulating variant (n = 217). Baseline values correspond to severe disease. Model specification: ln(1-VE) ~ (time since booster + time since immunogen emergence) * outcome.*

| Parameter | Estimate (95% CI) | p-value |
|---|---|---|
| Intercept | 0.2378 (0.1400, 0.4041) | <0.0001 |
| Time since booster administration (months) | 1.099 (1.076, 1.122) | <0.0001 |
| Time since immunogen emergence (months) | 1.023 (1.002, 1.044) | 0.0293 |
| Outcome=Symptomatic | 1.217 (1.098, 1.349) | 0.0002 |
| Outcome=Death | 0.6514 (0.5348, 0.7934) | <0.0001 |
| Mean/Median Age (years) | 1.003 (0.9965, 1.009) | 0.3785 |

*Supplementary Table 13 Results of combined meta-analytic model fitted to vaccine effectiveness data following vaccination with any immunogen. Note that baseline values are for severe disease. Data includes vaccine effectiveness estimates stratified by time since booster administration and circulating variant (n = 101). Baseline values correspond to severe disease. Model specification: ln(1-VE) ~ time since booster + time since immunogen emergence + outcome + age (median or mean).*

| Parameter | Estimate (95% CI) | p-value |
|---|---|---|
| Intercept | 0.2954 (0.2479, 0.3520) | <0.0001 |
| Time since booster administration (months) | 1.089 (1.072, 1.106) | <0.0001 |
| Time since immunogen emergence (months) | 1.023 (1.011, 1.036) | 2e-04 |
| Outcome=Symptomatic | 1.199 (1.122, 1.281) | <0.0001 |
| Outcome=Death | 0.8309 (0.7462, 0.9251) | 7e-04 |

*Supplementary Table 14 Results of combined meta-analytic model fitted to vaccine effectiveness data from cohorts involving participants over 18 years of age only following vaccination with any immunogen. Data includes vaccine effectiveness estimates stratified by time since booster administration and circulating variant (n = 209). Baseline values correspond to severe disease. Model specification: ln(1-VE) ~ time since booster + time since immunogen emergence + outcome.*

| Parameter | Estimate (95% CI) | p-value |
|---|---|---|
| Intercept | 0.2610 (0.2150, 0.3170) | <0.0001 |
| Time since booster administration (months) | 1.088 (1.072, 1.104) | <0.0001 |
| Time since immunogen emergence (months) | 1.027 (1.015, 1.040) | <0.0001 |
| Outcome=Symptomatic | 1.161 (1.091, 1.236) | <0.0001 |
| Outcome=Death | 0.8359 (0.7521, 0.9291) | 0.0009 |
| North America | 1.164 (1.024, 1.324) | 0.0204 |
| Asia Pacific | 1.031 (0.7463, 1.424) | 0.8537 |

*Supplementary Table 15 Results of combined meta-analytic model fitted to vaccine effectiveness data following vaccination with any immunogen. Data includes vaccine effectiveness estimates stratified by time since booster administration and circulating variant (n = 217). Baseline values correspond to severe disease in Europe. Model specification: ln(1-VE) ~ time since booster + time since immunogen emergence + outcome + study location.*

| Parameter | Estimate (95% CI) | p-value |
|---|---|---|
| Intercept | 0.3026 (0.2512, 0.3646) | <0.0001 |
| Time since booster administration (months) | 1.090 (1.074, 1.106) | <0.0001 |
| Time since immunogen emergence (months) | 1.023 (1.011, 1.035) | 0.0001 |
| Outcome=Symptomatic | 1.171 (1.100, 1.246) | <0.0001 |
| Outcome=Death | 0.8338 (0.7507, 0.9261) | 0.0007 |
| Other | 0.9599 (0.8435, 1.092) | 0.5351 |

*Supplementary Table 16 Results of combined meta-analytic model fitted to vaccine effectiveness data following vaccination with any immunogen. Data includes vaccine effectiveness estimates stratified by time since booster administration and circulating variant (n = 217). Baseline values correspond to severe disease in a test negative case-control study. Model specification: ln(1-VE) ~ time since booster + time since immunogen emergence + outcome + study type.*

| Parameter | Estimate (95% CI) | p-value |
|---|---|---|
| Intercept | 0.2623 (0.2076, 0.3314) | <0.0001 |
| Time since booster administration (months) | 1.087 (1.070, 1.104) | <0.0001 |
| Time since immunogen emergence (months) | 1.030 (1.015, 1.047) | 0.0002 |
| Outcome=Symptomatic | 1.172 (1.093, 1.255) | <0.0001 |
| Outcome=Death | 0.8286 (0.7454, 0.9211) | 0.0005 |
| JN.1 immunogen | 1.074 (0.9170, 1.257) | 0.3769 |
| KP.2 immunogen | 1.152 (0.9065, 1.465) | 0.2467 |

*Supplementary Table 17 Results of combined meta-analytic model fitted to vaccine effectiveness data following vaccination in cohorts given only a single booster immunogen. Data includes vaccine effectiveness estimates stratified by time since booster administration and circulating variant (n = 200). Baseline values correspond to severe disease after having received the XBB.1.5 booster immunogen. Model specification: ln(1-VE) ~ time since booster + time since immunogen emergence + outcome + booster immunogen.*

**Studies Identified from the ViewHub Systematic Review**


1. Abdul Aziz, N., Kirsebom, F.C.M., Allen, A. & Andrews, N. Effectiveness of spring 2024 (XBB.1.5) and autumn 2024 (JN.1) COVID-19 vaccination against hospitalisation in England. *Vaccine* **67**, 127870 (2025).
2. Andersen, K.M., *et al.* 2024–2025 BNT162b2 COVID-19 vaccine effectiveness in non-immunocompromised adults: mid-season estimates from vaccine registries in two states linked to administrative claims. *Vaccine* **62**, 127534 (2025).
3. Andersen, K.M., *et al.* Effectiveness of BNT162b2 XBB.1.5 vaccine in immunocompetent adults using tokenization in two U.S. states. *Vaccine* **52**, 126881 (2025).
4. Andersson, N.W., *et al.* Comparative effectiveness of monovalent XBB.1.5 containing covid-19 mRNA vaccines in Denmark, Finland, and Sweden: target trial emulation based on registry data. *BMJ Medicine* **3**, e001074 (2024).
5. Antunes, L., *et al.* Early COVID-19 XBB.1.5 Vaccine Effectiveness Against Hospitalisation Among Adults Targeted for Vaccination, VEBIS Hospital Network, Europe, October 2023–January 2024. *Influenza and Other Respiratory Viruses* **18**, e13360 (2024).
6. Caffrey, A.R., *et al.* Effectiveness of BNT162b2 XBB vaccine in the US Veterans Affairs Healthcare System. *Nature Communications* **15**, 9490 (2024).
7. Cai, M., Xie, Y. & Al-Aly, Z. Association of 2024–2025 Covid-19 Vaccine with Covid-19 Outcomes in U.S. Veterans. *New England Journal of Medicine* **393**, 1612–1623 (2025).
8. Carazo, S., *et al.* Monovalent mRNA XBB.1.5 vaccine effectiveness against COVID-19 hospitalization in Quebec, Canada: Impact of variant replacement and waning protection during 10-month follow-up. *PLOS ONE* **20**, e0325269 (2025).
9. Ciesla, A.A., *et al.* Effectiveness of 2023–2024 Coronavirus Disease 2019 (COVID-19) Vaccines in Pregnant Women. *Obstetrics & Gynecology* **147**, 569–573 (2025).
10. DeCuir, J., *et al.* Interim Effectiveness of Updated 2023-2024 (Monovalent XBB.1.5) COVID-19 Vaccines Against COVID-19-Associated Emergency Department and Urgent Care Encounters and Hospitalization Among Immunocompetent Adults Aged ≥18 Years - VISION and IVY Networks, September 2023-January 2024. *MMWR Morb Mortal Wkly Rep* **73**, 180–188 (2024).
11. Du, Y., Paritala, S., Xu, Y., Maloney, P. & Lin, D.-Y. Durability of 2024-2025 COVID-19 Vaccines Against JN.1 Subvariants. *JAMA Internal Medicine* **185**, 1501–1504 (2025).
12. Fotakis, E.A., *et al.* Impact of the 2023/24 autumn-winter COVID-19 seasonal booster campaign in preventing severe COVID-19 cases in Italy (October 2023–March 2024). *Vaccine* **42**, 126375 (2024).
13. Hansen, C.H., Lassaunière, R., Rasmussen, M., Moustsen-Helms, I.R. & Valentiner-Branth, P. Effectiveness of the BNT162b2 and mRNA-1273 JN.1-adapted vaccines against COVID-19-associated hospitalisation and death: a

Danish, nationwide, register-based, cohort study. *The Lancet Infectious Diseases* **25**, 1293–1302 (2025).

27. Liu, B., Scaria, A., Stepien, S. & Macartney, K. Effectiveness of COVID-19 vaccine boosters for reducing COVID-19 mortality among people aged 65 years or older, Australia, August 2023 – February 2024: a retrospective observational cohort study. *Medical Journal of Australia* **223**, 418–422 (2025).
28. Liu, B., Stepien, S. & Macartney, K. Changing impact of COVID-19 vaccination on COVID-19 mortality in the SARS-CoV-2 omicron variant period, 2022-2025. *medRxiv*, 2025.2011.2020.25340696 (2025).
29. Ma, K.C., *et al.* Effectiveness of Updated 2023–2024 (Monovalent XBB.1.5) COVID-19 Vaccination Against SARS-CoV-2 Omicron XBB and BA.2.86/JN.1 Lineage Hospitalization and a Comparison of Clinical Severity—IVY Network, 26 Hospitals, 18 October 2023–9 March 2024. *Clinical Infectious Diseases* **82**, e595–e603 (2026).
30. Ma, K.C., *et al.* Estimated Effectiveness of 2024-2025 COVID-19 Vaccination Against Severe COVID-19. *JAMA Network Open* **9**, e2557415–e2557415 (2026).
31. Merdrignac, L., *et al.* Effectiveness of XBB.1.5 Vaccines Against Symptomatic SARS-CoV-2 Infection in Older Adults During the JN.1 Lineage-Predominant Period, European VEBIS Primary Care Multicentre Study, 20 November 2023–1 March 2024. *Influenza and Other Respiratory Viruses* **18**, e70009 (2024).
32. Monge, S., *et al.* Effectiveness of XBB.1.5 Monovalent COVID-19 Vaccines During a Period of XBB.1.5 Dominance in EU/EEA Countries, October to November 2023: A VEBIS-EHR Network Study. *Influenza and Other Respiratory Viruses* **18**, e13292 (2024).
33. Nguyen, J.L., *et al.* Effectiveness of the BNT162b2 XBB.1.5-adapted vaccine against COVID-19 hospitalization related to the JN.1 variant in Europe: a test-negative case-control study using the id.DRIVE platform. *eClinicalMedicine* **79**(2025).
34. Nunes, B., *et al.* Monovalent XBB.1.5 COVID-19 vaccine effectiveness against hospitalisations and deaths during the Omicron BA.2.86/JN.1 period among older adults in seven European countries: A VEBIS-EHR network study. *Expert Review of Vaccines* **23**, 1085–1090 (2024).
35. Rojas-Benedicto, A., *et al.* Effectiveness of the autumnal COVID-19 vaccine dose during the winter and summer waves of the 2023/24 season in Spain. *Expert Review of Vaccines* **24**, 980–990 (2025).
36. Separovic, L., *et al.* Effectiveness of 2024/25 KP.2 vaccine against outpatient COVID-19 in Canada. *medRxiv*, 2025.2009.2005.25335203 (2025).
37. Tang, L., *et al.* WSK-V102C COVID-19 vaccine effectiveness against SARS-CoV-2 JN.1 symptomatic infection among adults 60 years and older: a prospective observational cohort study in Shaanxi Province, China, 2024. *BMC Public Health* **26**, 131 (2025).
38. Tartof, S.Y., *et al.* Estimated Effectiveness of the BNT162b2 XBB Vaccine Against COVID-19. *JAMA Internal Medicine* **184**, 932–940 (2024).
39. U. K. Health Security Agency. COVID-19 vaccine surveillance report: November 2024. (UK Health Security Agency, London, 2024).
40. Vicic, N., *et al.* Evaluating the Effectiveness of 2024–2025 Seasonal mRNA-1273 Vaccination Against COVID-19-Related Hospitalizations and Medically Attended COVID-19 Among Adults Aged ≥ 18 years in the United States: An Observational Matched Cohort Study. *Infectious Diseases and Therapy* **15**, 701–714 (2026).

41. Volkman, H.R., *et al.* Durability of the BNT162b2 XBB:1.5-adapted vaccine against JN.1 hospitalisation in Europe, October 2023 to August 2024: A test-negative case-control study using the id.DRIVE platform. *PLOS ONE* **21**, e0342382 (2026).
42. Ward, T., *et al.* Understanding the effectiveness of the Comirnaty monovalent and bivalent vaccines during the Winter Coronavirus (COVID-19) Infection Study. *Journal of Infection* **90**(2025).
43. Wilson, A., *et al.* Evaluating the Effectiveness of mRNA-1273.815 Against COVID-19 Hospitalization Among Adults Aged ≥ 18 Years in the United States. *Infectious Diseases and Therapy* **14**, 199–216 (2025).
44. Zheng, Z., *et al.* Effectiveness of 2023–2024 mRNA-1273 XBB.1.5 vaccine against COVID-19 associated hospitalizations and medically attended COVID-19 in the United States. *Vaccine: X* **27**, 100737 (2025).
